# Determinants associated with preterm births in Uganda: A cross-sectional study

**DOI:** 10.1101/2022.11.25.22282746

**Authors:** Bill Nkeeto, Bruno L. Yawe, Fred Matovu

## Abstract

**Background:** Preterm births affect households’ incomes through direct and indirect expenditures associated with low productivity and the actual loss of employment in many cases. We studied the determinants of preterm birth in Uganda as one of the major contributors to neonatal morbidity and mortality, leading to households’ economic losses.

**Methods:** We used a cross-sectional research design based on the most recent Uganda Demographic Health Survey of 2016. The sample contained 1,537 women aged 15-49 years. The variable selection process was guided by categorization of the variables into; socio-demographic, reproductive history, and gestational birth characteristics. The study adopted two means of analysis. The logistic regression model to determine variables of preterm birth between 22 – 36 weeks and normal delivery period. Then the multinomial logistic regression model to determine how two preterm birth categories (22 – 32 weeks and 33 – 36 weeks) relate with the normal delivery period.

**Results:** Belonging to the poorest quintile (AOR2.09, 95% CI (1.69-2.57)) and attending antenatal care less than four times (AOR1.41, 95% CI (1.20-1.66)) had the highest odds ratios for the logistic regression model. Whereas the multinomial logistic regression model; for the 22-32 weeks category, belonging to the poorest quintile (RRR2.43, 95% CI(1.45-4.08)), attending antenatal care less than four times (RRR2.44, 95% CI (1.63-3.64)), had the highest relative risk ratios. For the 33-36 weeks category; belonging to a poorest quintile (RRR2.03, 95% CI (1.62-2.53)), having had less than four antenatal visits (RRR1.29, 95% CI (1.09-1.54)), and unwanted pregnancy (RRR1.22, 95% CI (1.03-1.45)), had the highest relative risk ratios.

**Conclusion:** Attending antenatal care for less than four times and belonging to the poorest quintile are common risk factors related to preterm birth. We therefore recommend that these receive utmost attention from the policy makers and implementers.

## Introduction

Preterm birth (PTB) refers to the birth of children before 37 weeks of pregnancy (1). Preterm births are the third leading cause of newborn mortality in Uganda, after malaria and pneumonia (2). This situation explains why Uganda’s neonatal mortality has persisted at 27 per 1000 live births for over two decades (3,4). Moreover, 14% of all the births in Uganda are PTB making Uganda rank 13^th^ globally in PTB rates out of 184 countries of the World Health Organization (WHO) (5). This The annual global figure of PTB is 15 million and these figures are feared to be on a rise (6). Of the 15 million PTBs each year, one million die from circumstances associated with premature births (7). Challenges associated with preterm birth (PTB) do not only affect the health expenditure of households but disorient health budgets of nations, an indication of a problem that requires constant tracking to mitigate or manage if it is inevitable (8). Consequently, the annual global statistics related to PTB are such that for every 10 children born, one is a PTB, making it the main source of neonatal morbidity and mortality associated with 28 percent of neonatal deaths and 35 percent of under-five deaths globally (9). According to the study by Ferrero et al., in 2016, on cross-country individual participant analysis of 4.1 million singleton births in 5 countries with very high human development index, the year 2013 had PTB contribute the most number of under five deaths estimated at 6.3 million globally (6). This indicates that, 15.4% of the live births, followed by pneumonia at 14.9% and other birth-related complications at 10.5%. Consequently, the surviving neonates have costs associated with their health. The subsequent implication of having PTBs, is taking the biggest hospital budgets in terms of expenditure in the world. The big costs arise from among many requirements. The extended period spent by a PTB in hospitals, on average is twelve days more compared to a full-term delivered neonate. Besides, there are other medium-term and strategic challenges like persistent morbidity, among other adverse neonatal outcomes arising from giving birth to preterm neonates(10,11). Further evidence to this, is the estimated cost to the federal government of the United States of America, where premature births expenditure amounts to $26.6 billion annually; this is through lost productivity, educational expenditure, and health expenditure (10). Similarly, a prospective study carried out in the UK to ascertain the economic costs related to moderate PTB (32 – 33 weeks) and late PTB (34 – 36 weeks), established that in the first 24 months after birth; the average communal cost of taking care of a moderate PTB was £1232, whereas, the average communal cost of taking care of a late PTB was £1114 compared to a communal average cost of £132 for a neonate born at term (>37 weeks) (12). Moreover, developed countries like USA, UK among others have an average PTB prevalence of 9 percent, compared to low and middle income countries (LMIC) with an average PTB prevalence of 12 percent (13). This means that there are many health complications encroaching on health budgets of LMIC although they are not quantified and directly assigned to the PTB burden. If a country like Uganda lowers its percentage of PTB infants from 13.6 percent (7), to a figure below 10 percent, it can go a long way in reducing the expenditure on health-related expenses especially those stemming from PTB. This can be done through addressing and managing the common factors associated to preterm births. Additionally, the preterm birth assumed causes are linked to whether these births are spontaneous, or medically initiated due to inevitable pregnancy complications. Subsequently, the type of PTB is further linked to the economic status of a country with the PTB prevalence. Countries with higher incomes are associated with a higher prevalence of medically initiated PTBs. A study by Morisaki et al., in 2014 on risk factors for spontaneous and provider-initiated preterm delivery in high and low Human Development Index (HDI) countries, revealed a 20 percent provider-initiated PTB in countries (HDI) as compared to 40 percent in countries with high HDI(14). This implies that several complications which require medically initiated PTB in LMIC, go unattended to, contributing to the higher rates of maternal and child morbidity and mortality, given the complications that ensue. Additionally, PTB is generally associated with certain characteristics. Women with education less than primary level along with teenage mothers, were more susceptible to spontaneous PTB than medically initiated PTB, where mothers of more than 35 years were the main victims. More still, women with low Body Mass Index (BMI) and high BMI (obese), are also susceptible to medically initiated PTB. Besides, IVF pregnancies have higher odds of becoming PTB through medical initiation(14). There is also evidence from some studies indicating how very short birth intervals and again too long birth intervals, have a very high association with PTB (15). Therefore, this study sought to understand the determinants of preterm birth in Uganda using the 2016 Uganda Demographic Health Survey (UDHS).

## Methods

### Approvals

This study did not require formal approval except for the authorization acquired to use the secondary survey data from the Demographic Health Survey (DHS) Program via (http://www.dhsprogram.com/data/dataset_admin/login_main.cfm). The DHS officially obtained the Ugandan government approval and adhered to the ethical practices that included getting informed consent and guaranteeing confidentiality of the respondents.

### Study design, study population, and Sample selection

The study used a cross-sectional research design based on the most recent UDHS of 2016. Specifically, the 2016 UDHS gathered information on several key demographic indicators. However, this particular study focused on the sections of the UDHS related to PTB.

### Sample selection

There were 18506 women of ages 15 – 49 interviewed in the 2016 UDHS. From these 7961 women were dropped because of having never neither delivered a child nor having inadequate information required for this study. This left 10545 women with information about pregnancies. However, 9008 women were dropped because their pregnancies were carried to term. This left the sample of the study with 1537 singleton PTBs that were used as indicated in **fig1**.

**Figure 1:**
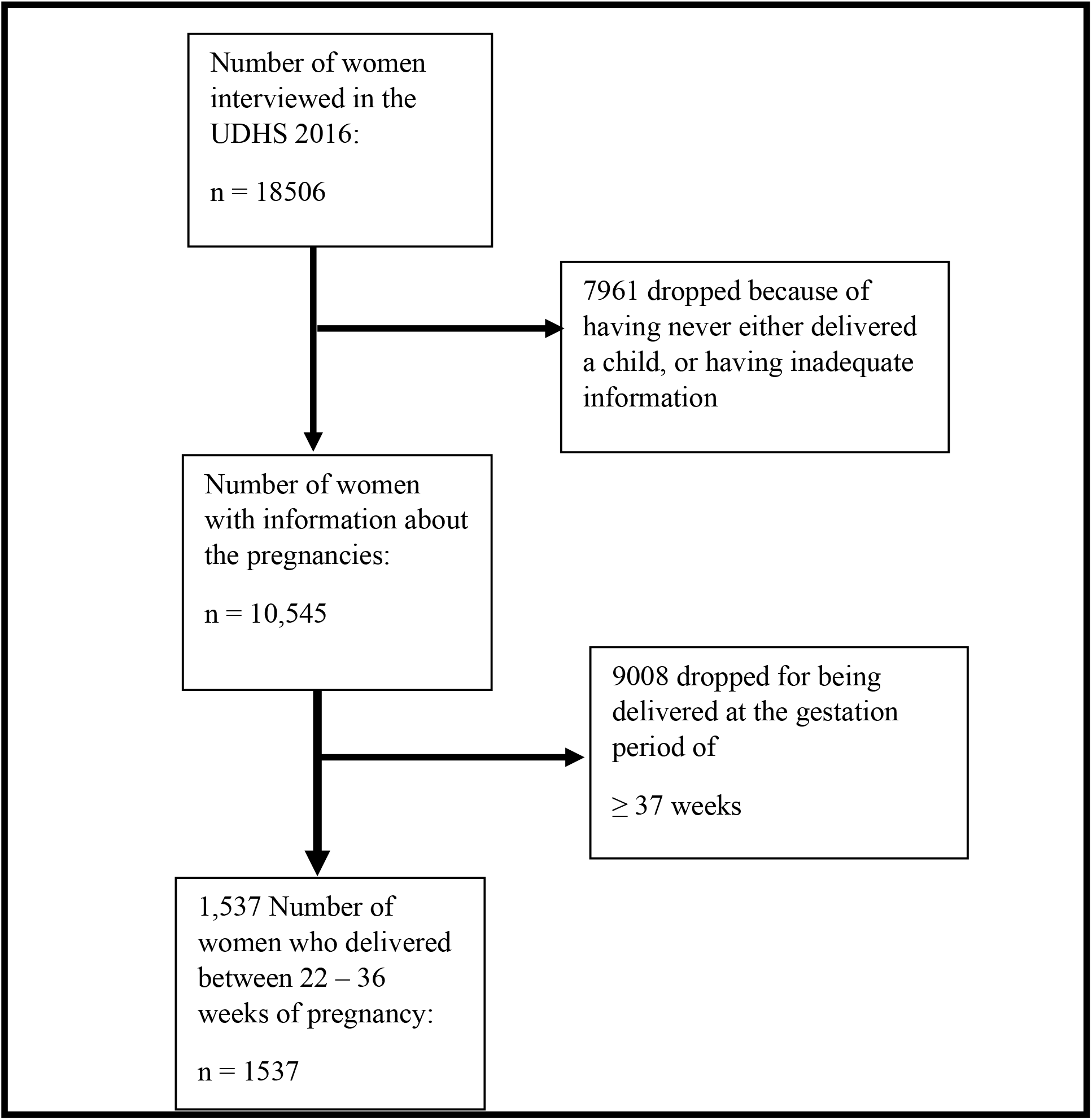
Sample selection criteria for determinants of preterm birth.

### Study variables

The study adopted two methods of analyzing the determining variables of preterm birth (PTB) in Uganda. The logistic regression model was used to determine PTB between 22 – 36 weeks and the neonates ≥37 weeks of birth. We further used the multinomial logistic regression (MLR) model to determine the association between PTB (22 – 32 weeks) and (33 – 36 weeks) with normal delivery (≥ 37 weeks) used as the reference category. The two methods of analysis were intended to establish whether the entire spectrum of PTB is influenced by similar factors or differently across the indicated PTB pregnancy gestation periods. This should hence guide the policy makers and decision making by practitioners faced with any PTB gestation category. Additionally, the study did not consider categorization of whether the PTB variables are spontaneously or medically initiated, since the data in the UDHS 2016 did not specify as such. Nevertheless, the variable selection process was guided by a model adopted from Medeiros et al., with modification of the contents, which categorizes the determinants into; socio demographic, reproductive history, and gestational birth determinants (13). We conceptualized the model by Medeiros in **fig 2** to cater for ogistic regression model and where **fig 3** was used to conceptualize the multinomial logistic regression.

**Fig 2.**
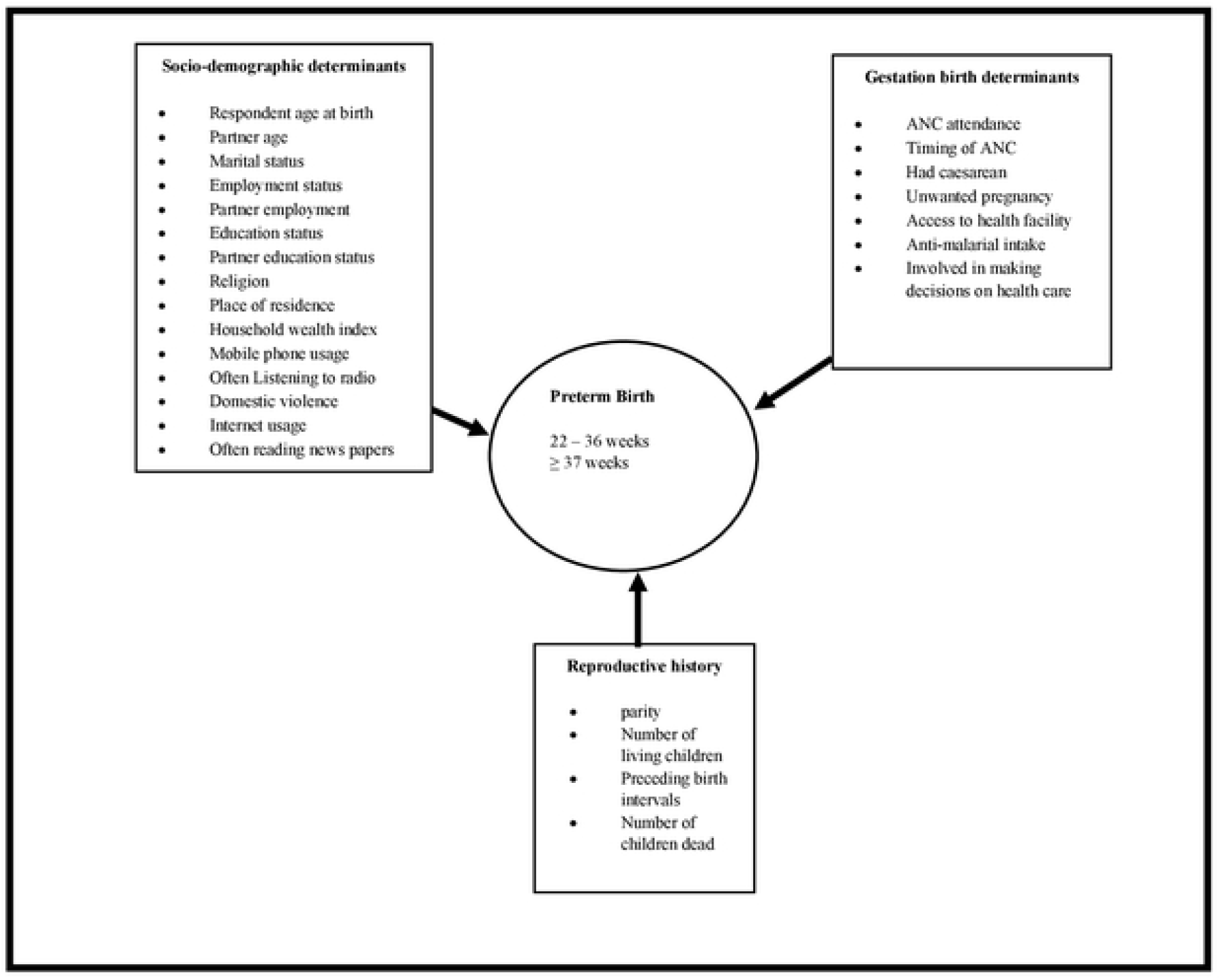
Conceptual framework for the determinants of birth weight (Logistic Regression model)

**Fig3-.**
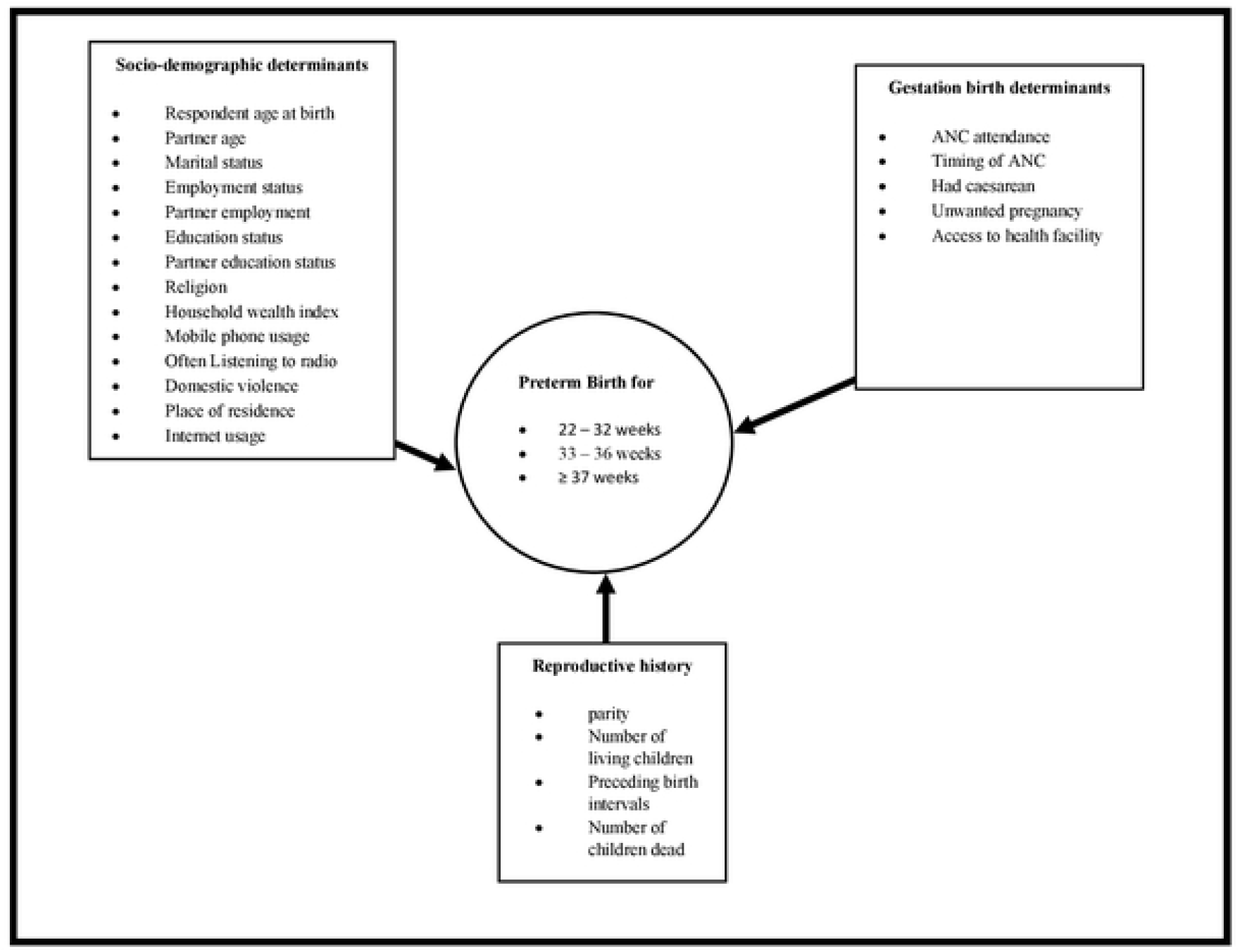
Conceptual framework for the determinants of birth weight (Multinomial Logistic Regression model)

Firstly, the socio-demographic variables included; religion, maternal education level, Age of the partner, maternal age at last birth, partner age, place of residence, respondent experienced domestic, respondents often listen to radio, respondent often uses mobile phone, respondent often reads newspapers, partner education level, household wealth index, marital status, employment status of the respondent, and partner employment status.

Secondly, the reproductive history determinants include the following; parity, preceding birth interval, number of dead children, and number of living children.

Thirdly, the gestation birth determinants included the following; antenatal care (ANC) attendance, ANC timing, access to the health facility, unwanted pregnancy, antimalarial intake, delivery by caesarean, and the respondent being involved in making health care decisions.

### Statistical analysis

This study had three forms of statistical analyses performed. That is, the summary statistics for both the dependent and independent variables were performed and presented. Then, bivariate analyses between the dependent variable and every each and every one of the independent variables were also performed ensuring that variables with a p ≤ 0.1 are taken to the next stage of analysis (16) except those backed by literature and logic to be of relevance in determining the dependent variable. Lastly, multivariate analyses were performed using a backward stepwise approach with the variables qualified from the bivariate stage.

Moreover, to ensure that the dependent variable gets the required explanations generically and more particularly at the selected PTB categories, this study adopted two models of analysis as already indicated. The study used the logistic regression (LR) model and the multinomial logistic (MLR) model. The results of this study at the multivariate stage of the LR model were reported using adjusted odds ratios. Whereas those of the MLR model, were reported using relative risk ratios (RRR). Subsequently, the level of statistical significance used was p ≤ 0.05. The goodness of fit of this study was determined by the Hosmer-Lemeshow test, for the LR model. Whereas the overall significance of the MLR model was determined by the likelihood ratio (LR) test.

Accordingly, the dependent variable with two values, 0 and 1 usually for No and Yes respectively, a logistic regression model is formed (17). For the first part of analysis of this study, the dependent variable is PTB = 1, otherwise = 0. Similarly, when the dependent variable has three or more unordered values, it is termed as multinomial logistic regression (17). For the second part of analysis for this study, the dependent variable is categorized into three categories; PTB of 22-32 weeks = 1, PTB of 33-36 = 2, and PTB of ≥37 weeks = 3.

For the logistic regression as required with the first part of analysis; if *p* is the proportion of the probability of giving birth to a PTB neonate which is given as 1 (one), then *1 - p* is the probability of not giving birth to a PTB neonate which is given as 0 (zero). According to (17), p**/**1-p indicates the odds and its logarithm indicates the log odds or the logit;

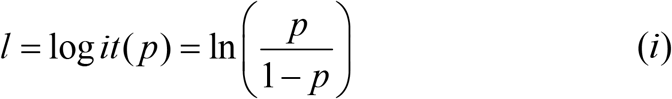

Subsequently, *p* ranges between 0 (zero) and 1 (one), whereas the logit can be anywhere in between minus (-) and plus (+) infinity. Moreover, the 0 (zero) logit happens with *p = 0*.*5*. So, creating a difference between two log-odds helps in comparing two probabilities, in this case, the probability of giving birth to PTB neonate *p*_1_ and the probability of not doing so *p*_2_ as may be demonstrated mathematically as;

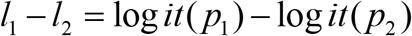

Taking logs gives;

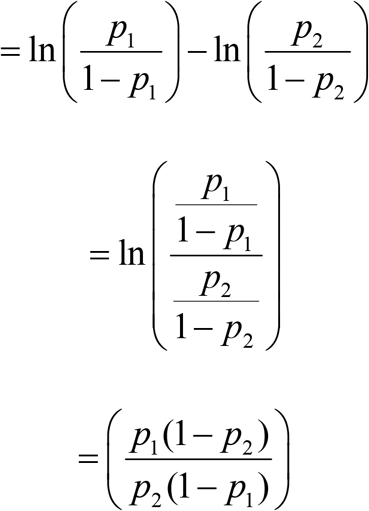

This gives the log odds ratio as;

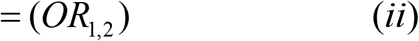

The odds ratio is instrumental during the comparison of probabilities across groups. It should also be noted that the logistic conversion is associated with the odds ratio; with the reverse association indicated as;

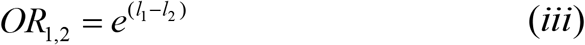

According to Badri, Toure and Lamontagne, a Logistic Regression (LR) is a modeling technique used to decide the association between a binary outcome variable and a set of *k* independent categorical or numerical variables [x_1_,x_2_,…,x_k_] (18). So, the occurrence or non-occurrence of an event *E* can be expressed as;

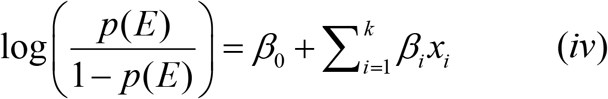

*β*_*1*_ represents the coefficients of the PTB independent determinants in the LR model, *β*_*0*_ is the intercept, while *p(E)* is the probability that an outcome variable is PTB determined by a given independent variable, for the LR bivariate model, or set of variables in a LR multivariate model. So equation *(iv)* will be used for the first part of this study, which requires the logistic regression model to establish which independent variables influence or do not influence PTB, from a list of variables.

Moreover, the multinomial logistic regression (MLR) model was used to establish the relation between several independent variables against the unordered categorical outcome variable PTB (22-32 weeks), PTB(33-36weeks), and PTB (≥37 weeks) which is a referent group. MLR permits, every grouping of an unordered response variable to be related to a reference category by providing various LR models (18).

The MLR model with the three categories indicated above, *k* categories of the outcome variable, the model consists of *k-1* concurrent logit equations as indicated below:

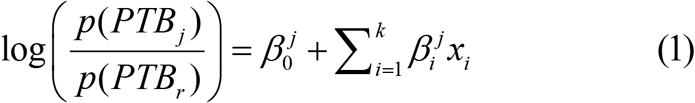

With *j = 1, …, k-1* where from equation *l, r* represents the reference category, in the case of this study, PTB ≥37 weeks’ category.

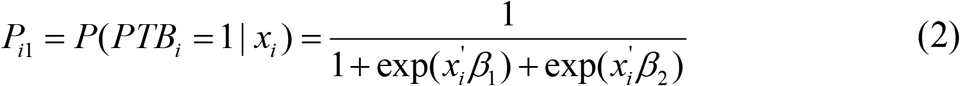

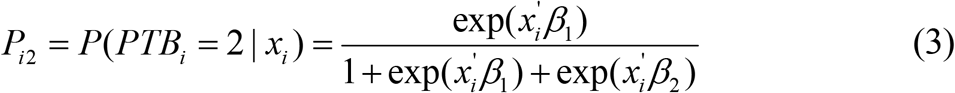

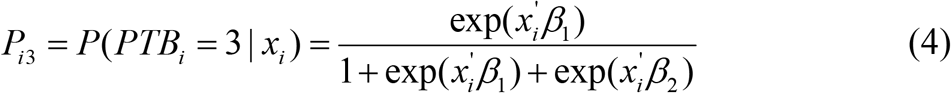

We made category 3 (≥37 weeks) of the dependent variable as the base category; so *β*_*1*_ and *β*_*2*_ indicate the covariate’s responses towards the first category (22-32 weeks of PTB) and the second category (33-36 weeks of PTB) respectively. Hosmaer and Lemeshow indicate that, for a three category dependent variable, any of the three can be determined to be a base category, that is, categories 1 and 2, to leave category 3 to be the base, or 2 and 3 to leave category 1 to be the base. This way, two logistic regression models are established (19). For this study, category 3 was used as the base to ensure that independent variables related to category 1 (22-33weeks) and those related to category 2 (33-36 weeks) are established with relation to it as the referent category (≥37 weeks).

Further, Hosmer, Lemeshow and Sturdivant indicate that the total of all the probabilities of the dependent variable categories, should sum up to 1 (one),(20) as shown below;

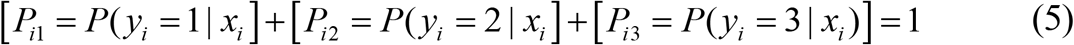

So the above probabilities of a dependent variable having *j* categories can be given in form of a multinomial logit as follows;

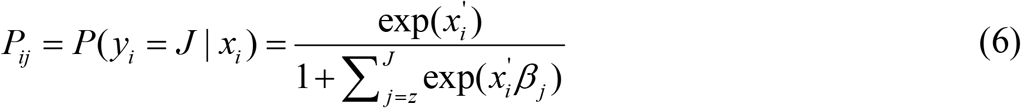

Moreover, for a single probability in category *j* of the dependent variable can be expressed as;

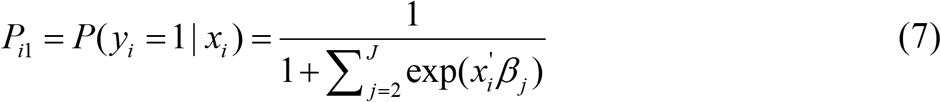

According to Ari and Aydin, to estimate the multinomial logit model, a maximum likelihood is used with the log-likelihood function for a given sample of *n* observations stated as;

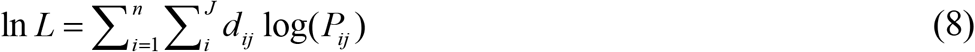

With *d*_*ij*_ denoting a dummy variable that takes a value of 1 (one) if observation *i* belongs to the *j*^*th*^ category of the dependent variable and 0 (zero) if otherwise, since *P*_*ij*_ is a non-linear function of the constraints of the model being regressed (21).

This study used relative risk ratio (RRR) to interpret the results of all significant variables. That is, the 22 – 32 weeks PTB category relative to the base category, ≥ 37 weeks PTB category. Similarly, the 33 – 36 weeks PTB category, relative to the base category ≥ 37 weeks PTB base category. The RRR is got by exponentiation of the multinomial logit coefficients, e^coef^ (22). This study, specified the RRR with a stata command of mlogit, RRR. Since the ML model estimates *k–1* models, where the *k*^*th*^ equation is relative to the base outcome. For this study, the RRR of a coefficient in a 22 – 32 weeks PTB category or 33 – 36 weeks PTB category, indicates how the risk of the outcome falling in any of the two categories, to the risk of the outcome falling in the ≥ 37 weeks PTB category. This changes with the variable in each of the two categories (22 – 32 weeks PTB category or 33 – 36 weeks PTB category). For a Relative risk ratio (RRR) > 1 shows that the risk of the outcome falling in the comparison category relative to the risk of the outcome falling in the base category increases as the variable increases. That is to say, the outcome is most likely to occur on the side of the comparison category. Whereas, a RRR < 1 shows that the risk of the outcome falling in the comparison category relative to the risk of the outcome falling in the base category reduces as the variable increases. This implies that the outcome is in favour of the base category (22). STATA 16 was used during the analysis of the data of this study.

## Results

The study started by indicating the categorization of each of the covariates to be used. It further proceeded by establishing the summary statistics of each of the covariates to be used for further analysis as shown in **Table 1**.

**Table 1: categorization of variables and their summary statistics**.

The study analysis went further to carry out bivariate analysis for the two adopted methods of analysis, that is, the bivariate logistic regression analysis indicated in **Table 2** and the bivariate multinomial logistic regression analysis, where for both cases, variables which had a p-value of ≤ 0.2 (16), were selected for further multivariate analysis.

**Table 2: Bivariate logistic regression results for the preterm birth**

### Qualified variables from the bivariate logistic regression

Variables that were not significant at the threshold of p ≥ 0.1 at the bivariate logistic regression stage were not included using a backward stepwise approach at the multivariate logistic regression stage except where previous literature and logic required so. The variables that were eliminated for not meeting the threshold required p-value included; domestic violence at p = 0.794, caesarean birth at p = 0.531, and ANC timing at p = 0.722. Similarly, covariates such as higher education for both the respondents and their partners at p = 0.113 and p = 0.866 respectively were also not included. Others eliminated included; not having attended ANC at p = 0.436, having a partner of age category 50 – 95 at p = 0.300 and belonging to a poorer wealth quintile at p = 0.421. Consequently, internet usage was not significant at p = 0.655, but given the influence internet has on the information flow, it was included at the LR multivariate stage. Primary education of the respondent was also not significant at p = 0.795. This was due to the importance linked to this stage of education as the basic stage expected for one to acquire knowledge about maternal health services. More so, maternal education commanded a 59% frequency response according to the summary statistics in **Table 1**. Primary education was included as one of the covariates under the respondents’ education status. The other covariate included at the multivariate LR stage under religion, was being a Muslim at p = 0.360 given the influence religion has on peoples lifestyles and as a result affecting the way they make health decisions.

### Multivariate logistic regression

Before proceeding with the multivariate logistic analysis, all the qualified variables were subjected to the collinearity test as indicated in **annexure 1**, using stata 16 command of the pairwise correlation (pwcorr). As such; parity was highly correlated with maternal age at birth with correlation coefficient r = 0.812. Nevertheless, both variables standout as key determinants of maternal and early childhood health, however, the issues associated with the average number of children born by a woman in the developing world are still encompassed with more risks compared to those surrounding maternal age (23). So, parity was chosen over maternal age during the LR multivariate analysis of this study.

### Results for the multivariate Logistic regression analysis for the determinants of preterm birth

Subsequently, the determinants from the bivariate LR stage included in the multivariate LR model, were selected using the backward stepwise approach. The results presented in **Table 3**, were published using a stata command asdoc formed by Shah, (2018); indicating their adjusted odds ratios, standard errors, t-values, p-values, and the 95% confidence intervals. Nevertheless, due to collinearity, the model automatically omitted eight (8) determinants, accordingly: higher education for both the respondents and their partners; first parity and 5 plus parity; marital status; being a rural resident; respondent being a casual employee; having a very old partner; and not having born children before. Besides, only significant variables were retained in **Table 3**.

**Table 3: Multivariate Logistic regression results on determinants of preterm birth**

After carrying out the multivariate LR analysis of the model, the testing of the overall significance of the model was done using the Hosmer-Lemeshow test with the stata command (estat gof, group (10) table) as shown in **Table 4**. This test showed that the overall fit of the model was good at; with 6643 number of observations, Hosmer-Lemeshow chi2 (8) = 8.75 and Prob > chi2 = 0.3634, as indicated in **Table 4**.

**Table 4:** Logistic model for premature, goodness-of-fit test.

| Group | Prob | Obs 1 | Exp 1 | Obs 0 | Exp 0 | Total |
| --- | --- | --- | --- | --- | --- | --- |
| 1 | 0.044 | 24 | 23.800 | 641 | 641.200 | 665 |
| 2 | 0.057 | 24 | 33.900 | 640 | 630.100 | 664 |
| 3 | 0.068 | 41 | 41.400 | 623 | 622.600 | 664 |
| 4 | 0.079 | 48 | 48.700 | 617 | 616.300 | 665 |
| 5 | 0.093 | 67 | 56.600 | 597 | 607.400 | 664 |
| 6 | 0.111 | 68 | 67.300 | 596 | 596.700 | 664 |
| 7 | 0.140 | 81 | 82.200 | 584 | 582.800 | 665 |
| 8 | 0.181 | 103 | 105.700 | 561 | 558.300 | 664 |
| 9 | 0.231 | 151 | 135.000 | 513 | 529.000 | 664 |
| 10 | 0.594 | 179 | 191.400 | 485 | 472.600 | 664 |

The output of the analysis in **Table 3** showed that the socio-demographic determinants category of the model, had two statistically significant variables at 1% (p ≤ 0.01) and 5% (p ≤ 0.05) respectively. The adjusted odds ratio (AOR) > 1 (risk factors) were; belonging to the poorest quintile with p = 0.000 at AOR = 2.1 and CI (1.692; 2.574). The other variable was internet usage at p = 0.042, AOR = 1.711 and CI (1.020; 2.870). Whereas the statistically significant variables at 1 % (p ≤ 0.01) and adjusted odds ratios (AOR) < 1 (protective factors) were; all the three partner education categories that were qualified to the multivariate LR stage were significant. Having an uneducated partner was significant at 1% with p = 0.000, with AOR = 0.450 and CI (0.303; 0.658). More so, having a partner who studied up to primary education level was significant at 1% with p = 0.000, AOR = 0.478 and CI (0.308; 0658). More still, having a partner at the secondary level of education was p = 0.006, AOR = 0.667 and CI (0.360; 0.892). Besides, belonging to a middle wealth quintile was significant at 5% with p = 0.029, AOR = 0.748 and CI (0.577; 0.970). Additionally, belonging to the richer wealth quintile was significant at 1% and had p = 0.003, AOR = 0.630, and CI (0.465; 0.835). Also, belonging to the richest wealth quintile, was significant at 1% with p = 0.005, AOR = 0.552, and CI (0.365; 0.835).

The other socio-demographic significant protective factors, included; residing in an urban area significant at 5% with p = 0.019, AOR = 0.712, and CI (0536; 0.945). Mobile phone usage significant at 5% with p = 0.029, AOR = 0.792, and CI (0.642; 0.976).

Furthermore, the gestation birth determinants only had two significant factors both risk factors. First, attending ANC for at least 2-3 times, significant at 1% with p = 0.000, AOR = 1.410 and CI (1.198; 1.660). Second, unwanted pregnancy, was significant at 5% with p = 0.025, AOR = 1.204, and CI (1024; 1.416).

Surprisingly, none of the reproductive history determinants was significant 5% at the multivariate stage except having had any dead child which was significant at 10% with p = 0.053 at AOR = 1.186. Equally from the socio-demographic factors, it was surprising not to have any of the respondents’ education level categories significant, as well as religion, given the influence the two variables have on health care decision making.

Additionally, as earlier indicated, the study also catered for the bivariate multinomial logistic regression analysis in **Table 5** where the variables which had a p-value of ≤ 0.2 (16), were selected for further multivariate analysis.

**Table 5: Bivariate Multinomial Logistic Model Results for PTB in Uganda**

### Interpretation of the results of the multivariate multinomial logistic regression analysis for determinants of preterm birth

All the variables in the study which had a p ≤ 0.1 as the threshold at the bivariate MLR stage for any of the two categories of preterm birth (22-32 weeks and 33-36 weeks, relative to ≥ 37 weeks), the outcome variable, were qualified to the multivariate MLR analysis stage. Others qualified included those backed by literature and logic, even when they were beyond the bivariate p-value set threshold. The covariates that were qualified even when they did not make the threshold p-value included; secondary education category of the partner and internet usage. Maternal age was not qualified since it was highly correlated with parity at r = 0.814, opting for parity given the maternal and childhood characteristics related to it (23). Conversely, covariates that were disqualified at the bivariate stage for not meeting the threshold p-value included; belonging to the catholic religious category. Even with the importance religion has, without the catholic religion, the model was having a better fit. Further, partner age of category 50-95 years, poorer quintile category, respondent belonging to the casual employment category, partner belonging to the casual employment category, having had caesarean delivery, not having attended ANC, and domestic violence, were also disqualified to the multivariate MLR model. The qualified covariates were further subjected to a backward stepwise approach to retain those that gave the best fit of the multivariate MLR model. These have their results indicated in **Table 6** developed using asdoc stata command created by Shah (2018). Consequently, due to collinearity, the model omitted; marital status, first parity, fifth parity, having had dead children, and having more than three living children. Besides, only significant variables were retained in **Table 6**.

To ensure that the model had a good overall fit, the study used “mlogitgof, group(10) table” stata command, suggested by Fagerland, Hosmer and Bofin in 2008 for testing the multinomial logistic regression models’ goodness of fit (25). The outcome revealed a good fit of the model with; 6643 number of observations, 3 outcome variables, with base outcome value 3, Chi2 statistic = 17.325 with 16 degrees of freedom and prob > Chi2 = 0.365, as indicated in **Table 7**.

**Table 7:** Table observed and expected frequencies.

| Group | Prob | Obs_3 Exp_3 | Obs_2 Exp_2 | Obs_1 Exp_1 | Total |
| --- | --- | --- | --- | --- | --- |
| 1 | 0.0439 | 642 641.78 | 18 20.42 | 5 2.79 | 665 |
| 2 | 0.0565 | 638 630.54 | 24 29.43 | 2 4.03 | 664 |
| 3 | 0.0675 | 623 622.90 | 36 35.95 | 5 5.15 | 664 |
| 4 | 0.0790 | 623 616.38 | 38 42.48 | 4 6.14 | 665 |
| 5 | 0.0931 | 588 607.01 | 67 49.77 | 9 7.22 | 664 |
| 6 | 0.1115 | 599 596.37 | 55 58.82 | 10 8.82 | 664 |
| 7 | 0.1399 | 587 582.17 | 68 71.76 | 10 11.07 | 665 |
| 8 | 0.1806 | 558 557.95 | 93 90.23 | 13 15.82 | 664 |
| 9 | 0.2313 | 514 528.66 | 122 112.88 | 28 22.45 | 664 |
| 10 | 0.5390 | 485 473.23 | 149 158.27 | 30 32.50 | 664 |

According to the results in **Table 5**, the socio-demographic factors, had belonging to the poorest quintile significant at 1% for the 22-32 weeks PTB category with a relative risk of 2.43 times relative to the base category of 37 plus weeks at p = 0.001, and CI (1.450; 4.084), holding all the other variables in the model constant. Mobile phone usage was significant at 5% would reduce the relative risk of belonging to the 22-32 weeks PTB category by 0.54 times relative to the referent group of 37 plus weeks, at p = 0.043 and CI(0.307;0.980); holding all other variables in the model constant.

Further, the gestation birth determinants related to the 22-32 weeks PTB category had having at least 2–3 ANC visits (less than four official visits), increase the relative risk of 2.44 times, give birth to a PTB of category 22–32 weeks category, relative to the reference category of more than 37 weeks significant at 1% (p = 0.000) and 95% CI (1.631; 3.640), holding other factors constant. Similarly, timing of the ANC within the first three months, would increase the relative risk of having a PTB within the 22-32 weeks, relative to the reference group of more than 37 weeks by 2.17 times, significant at 1% (p = 0.000) and CI(1.440; 3.280), holding other factors constant.

The PTB category of 33-36 weeks in **Table 5** had ten socio-demographic determinants and three gestation birth determinants significant. Accordingly, a unit increase with an unemployed partner, increases the relative risk of having a PTB neonate in the category of 33-36 weeks relative to the reference category of more than 37 weeks by 1.91 times significant at 1% (p = 0.000) and 95% CI(1.340; 2.744), holding other factors in the model constant. Surprisingly, all levels including having a partner who is not educated contribute toward reducing the relative risk of PTB in the category of 33-36 weeks. A unit increase in having an uneducated partner, reduces PTB of 33-36 weeks relative to the reference category of more than 37 weeks by 0.40 times significant at 1% (p = 0.000) and 95% CI (0.263; 0.597); holding other factors in the model constant. More so, a unit increase in a partner with primary education, reduces the relative risk of having a PTB baby in the 33-36 weeks’ category relative to the reference group category of more than 37 weeks by 0.48 times, significant at 1% (p = 0.000) and 95% CI (0.352; 0.650); holding other factors in the model constant. More still, having a unit increase in a partner with secondary education reduces the relative risk of having a PTB neonate in the 33 – 36 weeks category relative to the more than 37 weeks reference group by 0.65 times, significant at 1% (p = 0.006) with 95% CI(0.474; 0.884), holding other variables in the model constant.

Further with the socio-demographic factors are the household wealth index covariates. A unit increase in belonging to the poorest quintile, increases the relative risk of belonging to the 33-36 weeks PTB category relative to the more than 37 weeks reference group by 2.03 times, significant at 1% (p = 0.000) with 95% CI(1.624; 2.534), holding other variables in the model constant. Besides, a unit increase in the middle quintile reduces the relative risking of having a PTB neonate of 33-36 weeks category relative to the more than 37 weeks category by 0.73 times significant at 5% (p = 0.027) with 95% CI(0.551; 0.960), holding other factors in the model constant. Also, a unit increase in belonging to the richer quintile, reduces the relative risk of giving birth to a PTB neonate in a 33-36 weeks category relative to a more than 37 weeks referent category by 0.61 times significant at 1% (p = 0.002) with 95% CI(0.440; 0.840), holding other variables in the model constant. Lastly with the household wealth index is the unit increase in belonging to the richest quintile, reduces the relative risk of belonging to PTB category of 33–36 weeks relative to a more than 37 weeks referent category by 0.51 times significant at 1% (p = 0.003) with 95% CI(0.320; 0.790), holding other variables in the model constant. Conclusively with the socio-demographic factors, a unit increase in urban residence reduces the relative risk of having a PTB neonate in the category of 33–36 weeks category relative to a more than 37 weeks referent category by 0.69 times, significant at 5% (p = 0.015) with 95% CI(0.505; 0.930), holding other factors in the model constant. More so, significant at 10% (p=0.065) and 95% CI (0.692; 1.011) was belonging to an Anglican religion which reduces the relative risk of belong to the 33-36 weeks PTB category relate to the more than 37 reference category by 0.837 times, holding other factors in the model constant. Similarly, using of a Mobile phone reduces belonging to the 33-36 weeks PTB category by 0.827 times significant at 10% (p=0.094), holding other factors in the model constant.

Further still, are the gestation birth determinants having three significant variables and these include; attendance of 2 to 3 ANC times for which a unit increment, increases the relative risk of a PTB in the category of 33 – 36 weeks relative to the more than 37 weeks referent category by 1.30 times significant at 1% (p = 0.004) with CI (1.087; 1.540), holding other variables in the model constant. Moreover, a unit increase in unwanted pregnancy, increases the relative risk of giving birth to a PTB baby in the category of 33-36 weeks, relative to the more than 37 weeks referent category at by 1.22 times significant at 5% (p = 0.025) with 95% CI (1.090; 1.540), holding other factors in the model constant. To sum up the gestation birth determinants, a unit increase in the woman’s involvement in the health care decision making, reduces the relative risk of belonging to the PTB category of 33 – 36 weeks relative to the more than 37 weeks referent category by 0.82 times significant at 5% (p = 0.037) with 95% CI (0.682; 0.988), holding other factors in the model constant.

Finally, with the reproductive health characteristics, having had a dead child increases the chances of giving birth to 33-36 weeks PTB category neonate by1.181 times relative to the referent more than 37 weeks referent category, significant at 10% (p=0.077) with 95% CI(0.982;1.420), holding other factors in the model constant.

## Discussion

The WHO rates PTB as the main cause of neonatal mortality at 28% in Uganda (26). The most recent PTB figure as of 2016 is 1,665,000 births per year, of which 226,000 neonates are PTB from which 12,500 contribute toward Uganda’s child mortality of 15,464 (3,27). The study in the LRM revealed that the three significant covariates of partner education; no education, primary education, and secondary education are all associated with the reduction of PTB in this study. Primary education and secondary education are consistent with findings of studies by Dwarkanath et al., and Jungari and Paswan which found a positive association between spouse education and positive maternal and child health outcomes (28,29). Having an uneducated husband who contributes positively to positive health outcomes is in line with findings by Bhatta in Nepal, who however, finds these uneducated husbands to have high incomes that meet the necessities involved (30). The reduction seen in PTB arising from having uneducated husbands contributing positively to maternal and child health outcomes in Uganda, can be likened to the advocacy campaigns of male involvement by the MOH, among other players, and several social marketing activities that have created awareness. This has brought up a new wave of men that realize their mandate during maternal health care in spite of the setting in the health system being intimidating to some of them (31,32). Moreover, the MLRM did not find any husband education levels significantly related to the 22 – 32 weeks PTB category. However, 33 – 36 weeks PTB category in the MLRM was consistent with the findings of the LRM in view of the partner education related to preterm births.

For the household wealth with the LRM, save for the omitted poorer quintile due to collinearity, all the other four categories were significant with only the poorest quintile being associated with increased preterm birth. The rest of the wealth quintiles that were part of the multivariate LRM; is, middle quintile, richer quintile, and richest quintile; were associated with reduced PTB. This is in line with a study by Rai et al., West Bengal India that found increased wealth being positively associated with reduced PTB (33). Similarly, for the MLRM, for the PTB category of 22 – 32 weeks, it was on the poorest quintile that was significant with a unit increase in the poorest quintile leading to the relative risk of having PTBs. The rest of the three quintiles were consistent with the findings of the LRM. Similarly, for the category 33 – 36 weeks PTB category, only the poorest quintile indicated the increment in PTB. Whereas the middle quintile, richer quintile, and richest quintile; indicated reduced PTB. This implies poverty is one of the key causes of the very preterm births, indicated in both PTB categories of the MLRM as well as in the LRM. However, improved wellbeing is associated with reducing moderate PTB explained by 33 – 36 weeks PTB and/or general PTB reduction in the results from the LRM indicate. This implies that poverty reduction remains critical to enabling positive maternal and c outcomes (34,35).

Further, the LRM shows that staying in an urban area relative to a rural area, reduces PTB birth. This is based on the easy access to the readily available health facilities and the medical practitioners, which most rural areas in the country do not have. The MLR model also found the 33 – 36 weeks PTB category to, positively associated with being an urban resident reducing PTB in that category. Yet there was no significant association between the 22 – 32 Weeks category and PTB. This means that places of residence matter when the pregnancy goes into the 33 – 36 weeks. This shows an implied need for medical care that requires attention of several complications that arise during the 33-36 weeks period of the pregnancy. Similarly, extending better medical facilities and the human resources in the rural areas can be paramount in reduction of PTBs as well, for all the PTB categories. These findings are consistent with a study in Ethiopia by Abaraya and others and another study in the French Guiana by Leneuve-Dorilas and others, as well as consistent with findings of the study in Uganda on PTB by Ayebare others (7,36,37).

Furthermore, the study established with the LRM that having a mobile phone reduces the relative risk of having a PTB neonate. The results of MLRM however more specific on the 22 – 32 weeks PTB category. The mobile phone usage was not found to be a resourceful gadget for the 33 – 36 weeks PTB gadget. Subsequently, regardless of the category, the role played by the mobile phone in reduction of the adverse effects maternal and newborn outcomes is critical. There is evidence with previous studies that have tested usage of mobile phones and positive results in terms of awareness and increased visits of up to 43% for the mothers with signs of PTB registered (38).

The LRM also established that in Uganda, internet usage for the case of PTB had negative consequences for its increased usage. Results indicated that internet usage leads to increase in PTB. This can be attributed to the uncensored messages that are misinforming mothers of the proper ways to access and take action on matters related to PTB among other maternal and newborn health related issues. Besides, the inability to access the right gadgets to use and knowing how to use them to access relevant information using the internet, is a challenge to most mothers. This is in line with a study by Obasola and Mabawonku done in Nigeria that established usefulness of the information on maternal and child health distributed on the internet, with however, challenges in establishing how to access it (39). The MLRM results did not indicate a specific PTB category associated with internet to cause any relative effect to the PTB outcome. This implies that the effect of the internet to the PTB is generically affecting the entire PTB spectrum.

The gestation birth determinants; attending at least 2 - 3 ANC visits and timing of the first ANC visit being in the first three months. The LRM results revealed that doing 2 – 3 ANC visits and having the first ANC in the first three months was not sufficient enough to reduce the prevalence of PTB. These findings are consistent with the MLRM results which reveal a similar direction for the two PTB categories; 22 – 32 weeks and 33 – 36 weeks. The direction shown by these results is in line with the WHO new model of ANC which advocates for 8 visits other than four for the entire gestation period (40). This also implies that with more ANC visits in a shorter period of time, symptoms of PTB can be anticipated and mitigated. More so, the timing of the first ANC visit, was not significant with the LRM. However, timing of first ANC was significant with MLRM, specifically with the 22 – 32 weeks category where the current status of attendance indicates that expectant mothers do not attend the required number of times indicated by the WHO of between 8 and 12 weeks (41). This means that even when the number of times are done, but the timing is not right, there are consequences related to very preterm births which are likely to occur. Moreover, the new suggested frequency and time by WHO, that is, first contact in the first 12 weeks of gestation, and others to follow during the following weeks; 20, 26, 30, 34, 36, 38 and 40 weeks of gestation; can go a long way in sorting adverse maternal and newborn health adverse outcomes like preterm birth (41).

Besides, the LRM revealed that, unwanted pregnancy also increases the prevalence of PTB. Similarly, the MLRM revealed the same. However being specifically related to the 33 – 36 weeks PTB category. These findings are in line with the with what several studies have found out indicating how unwanted pregnancies tend to have their bearers not adhering to requirements like attending ANC, taking the proper medication, and getting the due care deserved (42,43). Nevertheless, the issues of unplanned pregnancies are common in Uganda, implying the PTB scare remains a reality unless more awareness on contraception and planning for the pregnancies is applied.

Lastly with the gestation birth determinants, is the involvement of birthing women in the health decision making. This variable was only significant with the MLRM with the 33 – 36 weeks PTB category where increase in women involvement in health care decision making reduces preterm birth by 0.82 times. These findings are in line with the study done in Nigeria which indicated how women involvement improved health care outcomes (44). Also consistent with another study done in Canada where women of pregnancies greater than 32 weeks were enrolled and gave good indication of what to be done when diagnosed with symptoms of preterm births (45). So promotion of women involvement as well as male involvement is critical in ensuring positive maternal and child health outcomes.

The LRM and MLRM results had the reproductive history variable, having had a dead child, increasing the risk of PTB for the MLRM and specifically for the 33-36 weeks category. This points to the need for extra care for mothers with a history of losing a newborn (46), especially with the week 33-36 weeks PTB category.

The strength of this study is the sample used which is representative enough to capture the entire country. The team which was engaged with the data collection and cleaning was highly trained to engage with the respondents in a highly professional and ethical manner. The variables that were collected information about were relevant for the information that was needed to carry out the investigation. The limitation of this study is mainly a response bias by the respondents. This is due to the respondents’ time-lag between giving birth and the time when the interview is conducted.

## Recommendation

The need to remain on track for achieving SDG 3.2 by 2030 is imperative for Uganda. This shall require reducing avoidable deaths of newborns and children under 5 years of age. This requires to reduce adverse outcomes like PTB that contribute towards neonatal death. Uganda will target to reduce neonatal death to 12 per 1,000 live births from the current 27 per 1000 live births; and under-5 mortality to 25 per 1,000 live births from the current 64 per 1000 live births by 2030 (47). This study establishes that; household poverty, low facilitation of health facilities in the rural areas, uncensored information used from the internet, low attendance of ANC, unwanted pregnancies are the risk factors associated with PTB. We recommend that there should be more sensitization on male involvement and the necessary roles to play. There should be adequate facilitation of health centers II, III and IV especially in rural areas. Further encourage the use of mobile phones for acquiring vital information from the health workers. Advocacy of increased use of contraceptives to reduce unwanted pregnancies. We also encourage equal decision making on health related issues for bother women and men in a household. There should be economic empowerment of households to reduce poverty which is related to adverse health outcomes like PTB.

## Data Availability

Data is attached

## Acknowledgments

We express appreciation to all participants of the UDHS 2016, Uganda Bureau of Statistics and ICF International. We also wish to thank Ester Lilian Acen for her in put during the final touches of putting together this manuscript.

**S1 Annexure: Pairwise correlation S2 Annexure: Datasets and Do-file**

**S2 Fig: Conceptual framework for the determinants of birth weight - Logistic Regression model**

**S3 Fig: Conceptual framework for the determinants of birth weight -Multinomial Logistic Regression model**

**S1 Table: Categorization of variables and their summary statistics**

**S2 Table: Bivariate logistic regression results for the determinants of preterm birth in Uganda**.

**S3 Table: Multivariate Logistic regression results for the determinants of preterm birth in Uganda**.

**S5 Table: Bivariate Multinomial Logistic Model Results for the determinants of preterm birth in Uganda**.

**S6 Table: Multivariate Multinomial Logistic Regression Results for the determinants of preterm birth in Uganda**.

## References

1. WHO. Preterm birth: Key facts. World Heal Organ. 2018;(February).

2. Waiswa P, Pariyo G, Kallander K, Akuze J, Namazzi G, Ekirapa-Kiracho E, et al. Effect of the Uganda Newborn Study on care-seeking and care practices: A cluster-randomised controlled trial. Glob Health Action. 2015;8(1).

3. UBOS & ICF International. UDHS 2016 final report [Internet]. Kampala; 2018 [cited 2018 Aug 19]. Available from: https://www.dhsprogram.com/pubs/pdf/FR333/FR333.pdf

4. MoH. Ministry of Health Strategic Plan 2020/21 - 2024/25. 2020. p. 1–188.

5. Nakubulwa C, Musiime V, Namiiro FB, Tumwine JK, Hongella C, Nyonyintono J, et al. Delayed initiation of enteral feeds is associated with postnatal growth failure among preterm infants managed at a rural hospital in Uganda. BMC Pediatr. 2020;20(1):1–9.

6. Ferrero DM, Larson J, Jacobsson B, Carlo G, Renzo D, Norman E, et al. Cross-Country Individual Participant Analysis of 4. 1 Million Singleton Births in 5 Countries with Very High Human Development Index Confirms Known Associations but Provides No Biologic Explanation for 2 / 3 of All Preterm Births. PLoS One. 2016;11(9):1–19.

7. Ayebare E, Ntuyo P, Malande OO, Nalwadda G. Maternal, reproductive and obstetric factors associated with preterm births in Mulago hospital, Kampala, Uganda: A case control study. Pan Afr Med J. 2018;30:1–8.

8. Vieira ACF, Alves CMC, Rodrigues VP, Ribeiro CCC, Gomes-Filho IS, Lopes FF. Oral, systemic and socioeconomic factors associated with preterm birth. Women and Birth [Internet]. 2019;32(1):e12–6. Available from: https://doi.org/10.1016/j.wombi.2018.02.007

9. Mekonen DG, Yismaw AE, Nigussie TS, Ambaw WM. Proportion of Preterm birth and associated factors among mothers who gave birth in Debretabor town health institutions, northwest, Ethiopia 11 Medical and Health Sciences 1114 Paediatrics and Reproductive Medicine. BMC Res Notes [Internet]. 2019;12(1):10–5. Available from: https://doi.org/10.1186/s13104-018-4037-7

10. Halimiasl A, Safari S, Hamrah MP. Epidemiology and related risk factors of preterm labor as an obstetrics emergency. Emergency. 2017;5(1):1–8.

11. Stylianou-Riga P, Kouis P, Kinni P, Rigas A, Papadouri T, Yiallouros PK, et al. Maternal socioeconomic factors and the risk of premature birth and low birth weight in Cyprus: A case-control study 11 Medical and Health Sciences 1117 Public Health and Health Services 11 Medical and Health Sciences 1114 Paediatrics and Reproductive Medicine. Reprod Health. 2018 Sep 19;15(1).

12. Khan KA, Petrou S, Dritsaki M, Johnson SJ, Manktelow B, Draper ES, et al. Economic costs associated with moderate and late preterm birth: A prospective population-based study. BJOG An Int J Obstet Gynaecol. 2015;122(11):1495–505.

13. Medeiros, Cornetta, Crispim, Cobucc. Obstetrics and Gynaecology Cases - Reviews Risk Factors Associated with Preterm Birth in a Brazilian Maternal and Child Health Hospital. Clin Med Int Libr. 2018;5(6):136.

14. Morisaki N, Togoobaatar G, Vogel JP, Souza JP, Rowland Hogue CJ, Jayaratne K, et al. Risk factors for spontaneous and provider-initiated preterm delivery in high and low Human Development Index countries: a secondary analysis of the World Health Organization Multicountry Survey on Maternal and Newborn Health. BJOG. 2014;121 Suppl:101–9.

15. Koullali B, Oudijk MA, Nijman TAJ, Mol BWJ, Pajkrt E. Risk assessment and management to prevent preterm birth. Semin Fetal Neonatal Med [Internet]. 2016;21(2):80–8. Available from: http://dx.doi.org/10.1016/j.siny.2016.01.005

16. Hilbe JM. Logistic Regression Models: Texts in Statistical Science [Internet]. Carlin BP, Faraway JJ, Tanner M, Zidek J, editors. Taylor & Francis Group; 2009. 73–75 p. Available from: http://www.crcpress.com

17. Statistical N, Ncss S. Logistic Regression. In: NCSS Statistcal software. p. 1–69.

18. Badri M, Toure F, Lamontagne L. Predicting unit testing effort levels of classes: An exploratory study based on Multinomial Logistic Regression modeling. Procedia Comput Sci [Internet]. 2015;62(Scse):529–38. Available from: http://dx.doi.org/10.1016/j.procs.2015.08.528

19. Hosmer DW, Lemeshow S. Applied Logistic Regression, second ed. John Wiley & Sons, Inc., Hoboken, New Jersey, USA. second edi. JOHN WILEY & Sons Ltd; 2000.

20. Hosmer D, Lemeshow S, Sturdivant RX. Applied Logistic Regression. [Internet]. Third Edit. Vol. 47, Biometrics. JOHN WILEY & Sons; 2013. 1632 p. Available from: https://onlinelibrary.wiley.com/doi/pdf/10.1002/9781118548387.fmatter

21. Ari E, Aydin N. Examination By Multinomial Logistic Regression Model Of The Factors Affecting The Types Of Domestic Violence Against Women : A Case Of Turkey. Int J Sci Technol Res. 2016;5(11).

22. Kwak C, Clayton-Matthews A. Multinomial logistic regression. Nurs Res. 2002;51(6):404–10.

23. Ndiaye K, Portillo E, Ouedraogo D, Mobley A, Babalola S. High-risk advanced maternal age and high parity pregnancy: Tackling a neglected need through formative research and action. Glob Heal Sci Pract. 2018;6(2):370–80.

24. Shah A. ASDOC: Stata module to create high-quality tables in MS Word from Stata output. Statistical Software Components S458466, Boston College Department of Economics. 2018.

25. Joseph Newton EH, Cox NJ, Bellocco R, Institutet K, Buis ML, Colin Cameron A, et al. The Stata Journal. Stata J [Internet]. 2012 [cited 2018 Jul 25]; Available from: http://www.stata-journal.comhttp//www.stata.com/bookstore/sj.htmlhttp://www.stata.com/bookstore/sjj.html http://www.stata-journal.com/archives.html

26. Bater J, Lauer JM, Ghosh S, Webb P, Agaba E, Bashaasha B, et al. Predictors of low birth weight and preterm birth in rural Uganda: Findings from a birth cohort study. PLoS One. 2020 Jul 1;15(7 July).

27. Preterm Birth Initiative (PTBi) Background. [cited 2021 Sep 15]; Available from: www.hppm.musph.ac.ug

28. Dwarkanath P, Vasudevan A, Thomas T, Anand SS, Desai D, Gupta M, et al. Socioeconomic, environmental and nutritional characteristics of urban and rural South Indian women in early pregnancy : fi ndings from the South Asian Birth Cohort (START) Public Health Nutrition. 2018;(3).

29. Jungari S, Paswan B. What he knows about her and how it affects her? Husband’s knowledge of pregnancy complications and maternal health care utilization among tribal population in Maharashtra, India. BMC Pregnancy Childbirth. 2019;19(1):1–12.

30. Bhatta DN. Involvement of males in antenatal care, birth preparedness, exclusive breast feeding and immunizations for children in Kathmandu, Nepal. BMC Pregnancy Childbirth [Internet]. 2013;13(14):1–10. Available from: http://www.biomedcentral.com/1471-2393/13/14

31. Kaye DK, Kakaire O, Nakimuli A, Osinde MO, Mbalinda SN, Kakande N. Male involvement during pregnancy and childbirth: Men’s perceptions, practices and experiences during the care for women who developed childbirth complications in Mulago Hospital, Uganda. BMC Pregnancy Childbirth. 2014;14(1):1–8.

32. Stern E, Pascoe L, Shand T, Richmond S. Lessons learned from engaging men in sexual and reproductive health as clients, partners and advocates of change in the Hoima district of Uganda. Cult Heal Sex [Internet]. 2015;17:190–205. Available from: http://dx.doi.org/10.1080/13691058.2015.1027878

33. Rai RK, Sudfeld CR, Barik A, Fawzi WW, Chowdhury A. Sociodemographic Determinants of Preterm Birth and Small for Gestational Age in Rural West Bengal, India. J Trop Pediatr. 2019;65(6):537–46.

34. Sadovsky ADI de, Matijasevich A, Santos IS, Barros FC, Miranda AE, Silveira MF. Socioeconomic inequality in preterm birth in four Brazilian birth cohort studies. J Pediatr (Rio J) [Internet]. 2018;94(1):15–22. Available from: http://dx.doi.org/10.1016/j.jped.2017.02.003

35. Snelgrove JW, Murphy KE. Preterm birth and social inequality : assessing the effects of material and psychosocial disadvantage in a UK birth cohort. ACTA Obstet Gynecol AOGS. 2015;94:766–75.

36. Abaraya M, Seid S, Ibro S. Determinants of preterm birth at Jimma University Medical Center, southwest Ethiopia. Pediatr Heal Med Ther. 2018;Volume 9:101–7.

37. Leneuve-Dorilas M, Favre A, Carles G, Louis A, Nacher M. Risk factors for premature birth in French Guiana: the importance of reducing health inequalities. J Matern Neonatal Med. 2019;32(8):1388–96.

38. Olivia Kim U, Barnekow K, Ahamed SI, Dreier S, Jones C, Taylor M, et al. Smartphone-based prenatal education for parents with preterm birth risk factors. Patient Educ Couns [Internet]. 2019;102(4):701–8. Available from: https://doi.org/10.1016/j.pec.2018.10.024

39. Obasola OI, Mabawonku IM. Mothers’ perception of maternal and child health information disseminated via different modes of ICT in Nigeria. Health Info Libr J. 2018;35(4):309–18.

40. WHO. WHO recommendation on antenatal care contact schedules [Internet]. 2018. Available from: https://extranet.who.int/rhl/topics/improving-health-system-performance/who-recommendation-antenatal-care-contact-schedules

41. WHO. WHO, Sexual and reproductive health New guidelines on antenatal care for a positive pregnancy experience. Geneva; 2016.

42. Sharifi N, Dolatian M, Kazemi AFN, Pakzad R. The relationship between the social determinants of health and preterm birth in Iran based on the WHO model: A systematic review and meta-analysis. Int J Women’s Heal Reprod Sci [Internet]. 2018;6(2):113–22. Available from: http://dx.doi.org/10.15296/ijwhr.2018.19

43. Goossens J, Branden Y Van Den, Sluys L Van Der, Delbaere I, Hecke A Van, Verhaeghe S, et al. The prevalence of unplanned pregnancy ending in birth, associated factors, and health outcomes. Hum Reprod. 2016;31(12):2821–33.

44. Osamor P, Grady C. Decision making autonomy:emperical evidence from Nigeria. J Biosoc Sci. 2018;50(1):70–85.

45. Ha V, McDonald SD. Pregnant women’ s preferences for and concerns about preterm birth prevention : a cross-sectional survey. BMC Pregnancy Childbirth [Internet]. 2017;17(49):1–10. Available from: http://dx.doi.org/10.1186/s12884-017-1221-z

46. Wolke D. Preterm birth: high vulnerability and no resiliency? Reflections on van Lieshout et al. (2018). J Child Psychol Psychiatry Allied Discip. 2018;59(11):1201–4.

47. SDSN. Indicators and a Monitoring Framework:Launching a data revolution for the Sustainable Development Goals [Internet]. [cited 2021 Aug 11]. Available from: https://indicators.report/targets/3-2/

